# Clinical and cost-effectiveness of diverse post-hospitalisation pathways for COVID-19: A UK evaluation utilising the PHOSP-COVID cohort

**DOI:** 10.1101/2024.07.15.24310151

**Authors:** the PHOSP-COVID Study Collaborative Group, AH Briggs, A Ibbetson, A Walters, L Houchen-Wolloff, N Armstrong, T Emerson, R Gill, C Hastie, P Little, C Overton, J Pimm, K Poinasamy, SJ Singh, S Walker, OC Leavy, M Richardson, O Elneima, HJC McAuley, A Shikotra, A Singapuri, M Sereno, RM Saunders, VC Harris, NJ Greening, EM Harrison, AB Docherty, NI Lone, JK Quint, JD Chalmers, L-P Ho, A Horsley, B Raman, LV Wain, CE Brightling, M Marks, RA Evans

## Abstract

**Background:** Long Covid has emerged as a complex health condition for millions of people worldwide following the COVID-19 pandemic. Previously, we have categorised healthcare pathways for patients after discharge from hospital with COVID-19 across 45 UK sites. The aim of this work was to estimate the clinical and cost-effectiveness of these pathways.

**Methods:** We examined prospectively collected data from 1,013 patients at 12-months post-discharge on whether they felt fully recovered (self-report), number of newly diagnosed conditions (NDC), quality of life (EQ-5D-5L utility score compared to pre-covid estimate) and healthcare resource costs (healthcare records). An analysis of the cost-effectiveness was performed by combining the healthcare resource cost and one-year EQ5D (giving a quality adjusted life-year: QALY) using statistical models that accounted for observed confounding.

**Results:** At 1 year, 29% of participants felt fully recovered and 41% of patients had an NDC. The most comprehensive services, where all patients could potentially access assessment, rehabilitation, and mental health services, were more clinically effective when compared with either no service or light touch services (mean (SE) QALY 0.789 (0.012) vs 0.725 (0.026)), with an estimated cost per QALY of £1,700 (95% uncertainty interval: dominated to £24,800).

**Conclusion:** Our analysis supports the need for proactive, stratified, comprehensive follow-up for adults after hospitalisation with COVID-19 showing these services are likely to be both clinically and cost-effective according to commonly accepted thresholds.

## Introduction

Long Covid remains a recognised ongoing health crisis. Despite the burden of disease there is a limited evidence base to guide service models, diagnostic modalities, and therapeutic interventions. Clinical care has evolved through expert opinion and experiential learning, with best practice advice and guidelines developed alongside (1)(2)(3). During the first year of the COVID-19 pandemic, healthcare pathways post-hospitalisation for patients with severe COVID-19 were based on hospital teams making their own judgements about what follow-up they would provide and to which patients (4). In Oct 2020 in England, UK, a national Long Covid Taskforce formed which included funding for specialist Long Covid clinics and a service specification was developed (5). To date there is minimal published research on what Long Covid services were set up internationally (6). The evidence from this scoping review recommended that most Long Covid healthcare should be situated in primary care and patients with complex symptoms should be referred to specialist Long Covid outpatient clinics, and depending on the patients’ needs, further referral to services such as rehabilitation should be considered.

Patients recovering from COVID-19 may experience new or worsening chronic conditions, for example diabetes, cardiac disease, anxiety, depression as well as ongoing symptoms in the absence of a defined chronic condition (Long Covid). As such, Long Covid can be a complex multifactorial condition and the UK National Institute for Clinical Excellence (NICE) recommends the availability of integrated multidisciplinary rehabilitation services for complex cases (2). Emerging evidence in community observational studies suggest that Long Covid is associated with increased health service resource use (7) and decreased quality of life (8). However, data on the effectiveness of rehabilitation in patients with Long Covid is limited (9). Most initial studies to date have been observational cohorts with no control group (10)(11)(12)(13) which cannot account for natural recovery whilst most randomised controlled trials are too small to be informative (14). The largest randomised controlled trial to date demonstrates benefit of a remotely delivered supervised programme (9) for patients post-hospitalisation and results are awaited for a face to face programme (15).

We previously described and categorised healthcare pathways created for patients after discharge from hospital with COVID-19 at 45 hospital sites across the UK participating in the PHOSP-COVID study at the time (16)(17). This classification included whether there was a service available or not, and the level of complexity and/or comprehensiveness of service provided assessed by four components: 1) which patients could access the service, for example, all patients versus only a sub-group such as only those who had received mechanical ventilation; 2) the level and complexity of the assessment; 3) the comprehensiveness of the rehabilitation service available; and 4) the comprehensiveness of the mental health services on offer. For the assessment, comprehensiveness was determined by the availability of a face-to-face assessment, use of a multi-disciplinary team, multi-system approach and the availability of complex diagnostics. Higher comprehensiveness/complexity of the rehabilitation and mental health interventions included in the service required a multi-dimensional holistic approach.

It is currently unclear how to optimally implement and stratify follow-up services to be holistic, integrated, equitable and both clinically and cost-effective. Understanding how to optimise healthcare support for individuals after severe COVID-19 to maximise quality of life and deliver services which are cost-effective is critical to personalised, high quality, value for money, care. The latter was highlighted as a priority question by patients and clinicians. (18) We therefore aimed to estimate the clinical and cost-effectiveness of identified pathways of post-hospitalisation care available during the first year of the COVID-19 pandemic.

## Methods

### PHOSP-COVID data set

We used data from the UK-based Post-HOSPitalisation COVID-19 (PHOSP-COVID) cohort study (19). Participants were recruited from hospitals across the UK, having been discharged between February 2020 and March 2021 with a discharge diagnosis confirming, or a suspected illness caused by, COVID-19. Only participants from the sites where the health services survey was completed so the healthcare pathway could be mapped were used (34/45 tier 2 sites) (16). A variety of data were assessed, alongside detailed holistic and multi-system assessments measured during participant follow-up visits reported in detail elsewhere (18). The data included: information relevant to the patient’s index admission, such as level of respiratory support, baseline health and demographics, Health Related Quality of Life (HRQoL) as measured by EQ5D instrument (20); known comorbidities at hospital admission (including: cardiac; respiratory; gastrointestinal; neurological and psychiatric; rheumatological; metabolic/endocrine/renal; and malignancy/haematological), and information related to use of health care resources. Participants were also asked to retrospectively complete the EQ5D-5L at the five-month visit estimating how they felt before their hospital admission for COVID-19 (pre-Covid).

In order to describe the post-covid sequelae of this population participants were asked whether they felt fully recovered from COVID-19 at around 5 and 12-month after discharge from hospital (available responses were yes, no or unsure) and Newly Diagnosed Conditions (NDC) were described. Indicators for NDCs were constructed from the data as conditions that were unrecorded prior to the hospital admission from COVID-19 and had a relevant objective investigation that was abnormal at one year post assessment: eGFR < 60 ml/min/1.73 m^2^ in patients without a previous diagnosis of chronic kidney disease; HbA1c >= 6.0% in patients without a previous diagnosis of diabetes; PHQ-9 >= 10 or GAD7 > 8 in patients without a previous diagnosis of depression or anxiety; MoCA < 23 in patients without a previous diagnosis of dementia; NTproBNP >= 400 ng/L or BNP >= 100 ng/L in patients without a previous diagnosis of heart failure.

#### EQ5D data and QALYs

We used the EQ5D-5L version of the EQ5D descriptive system to measure patient HRQoL (21). The survey assesses HRQoL across five dimensions: mobility, self-care, usual activities, pain/discomfort and anxiety/depression. Each dimension has 5 levels: no problems, slight problems, moderate problems, severe problems and extreme problems. Responses across the dimensions can be combined to give an overall utility index score which summarises the patient’s HRQoL.

In line with UK NICE recommendations, we mapped EQ5D-5L utility index scores to the 3-level version of the score (22). The utility scores collected in PHOSP-COVID were employed to estimate the resulting quality-adjusted life-years (QALYs) for the first-year post hospital discharge based on the modelled analysis of EQ-5D outcomes.

#### Healthcare resource data and associated costs

To estimate patient healthcare resource use we used self-report and available healthcare record data on primary, secondary and emergency care visits and medical investigations and procedures collected from bespoke clinical research forms at the two research visits. Unit cost data from the Health and Social Care Unit Cost database (23), National Schedule of NHS Costs (24) and Schedule of Events Cost Attribution Template (25) were used to estimate the costs associated with healthcare resource use. Resource use items available in PHOSP-COVID and the derived unit costs used in the analysis are summarised in Table S1 of Appendix C in the supplementary materials.

#### Healthcare pathways

Based on the previously reported typology (16) we utilised four indicator variables of whether the comprehensiveness of assessment of post-hospitalisation covid services was low/high, whether the comprehensiveness of rehabilitation services was low/high, whether the comprehensiveness of mental health services was low/high and whether services were available for all patients or targeted only at a sub-group of patients. Together these four variables described 16 possible permutations of the health care pathway, of which 11 unique pathways were represented within the 45 sites of the PHOSP-COVID study. Those site reporting ‘no service’ were considered to fall in to the ‘low’ category of all four variables in the typology.

### Statistical analysis of PHOSP-COVID data

We aimed to adjust for observed case-mix variables in our estimation of the potential impacts of health care pathways on EQ5D HRQoL / QALYs and Health Service Resource Costs. Available demographic, clinical and comorbidity data were used in a regression framework to estimate adjusted impacts of the four health care pathway variables described above. These were included into the regression equations as main effect variables, meaning that the 11 represented pathways in the PHOSP-COVID dataset were estimated by combining these four main effect variables estimated in the regression equations. Further detail of the precise form of these statistical models are presented in Appendix B of the supplementary materials. Alternative specifications of the models presented were explored, including using non recovery and existence of NDCs as mediating variables and as control variables.

#### Approach to Missing Data and Representativeness

In addition to the complete case analysis, we also undertook a two-step inverse probability weighting (IPW) analysis to reduce bias from both missing data and to account for selection bias of the PHOSP-COVID cohort compared to the more representative ISARIC study (26); the methodology has been previously described (27). IPW can correct the potential bias and improve the representativeness compared to complete case analysis, although it is generally less efficient statistically than multiple imputation for handling missingness that is assumed to be missing at random (28). Nevertheless, it has the advantage there is no need to create multiple complete data sets for analysis and so it is more efficient at a practical level.

### Estimation of cost-effectiveness of health care pathways

An analysis of the cost-effectiveness of the health care pathways seen in the PHOSP-COVID study was performed by combining the statistical equations for healthcare resource cost and one-year EQ5D (giving a quality adjusted life-year) for the different permutations of health care pathway offered, while holding all other variables in the regression constant at their mean values. Cost and QALYs for the different pathways identified are plotted on the cost-effectiveness plane with uncertainty represented by probabilistic sensitivity analysis (29).

## Results

### Statistical analysis of PHOSP data

#### Descriptives

Out of 2,697 Tier 2 study participants, there were 2,422 participants that were discharged from one of the 34 Tier 2 sites that provided data allowing their health care pathways to be mapped, and 2,100 had a one-year visit. Overall, 1,013 participants were included in the analysis sample with complete data for all variables including all the patient reported outcome measures, the assessments for the NDCs and their summary demographic and baseline clinical information. There was good concordance in terms of baseline characteristics for the full sample and the analysis sample (Table 1). Most patients in the analysis sample were male (62%), white (79%), aged ≥ 50 years (81%) and had a BMI ≥ 30 kg/m2 (58%). The most common comorbidities at baseline were cardiac (46%), respiratory (28%) and neurological and/or psychiatric (19%). WHO class 5 (supplemental oxygen (WHO, 2020)) was the most common level of respiratory support provided during hospitalisation (42.3%). There were relatively equal numbers of patients across quintiles of social deprivation (18.1% to 21.4%).

**Table 1.** Baseline demographic and clinical characteristics of analysis sample* and all those from sites with health care pathways mapped.

| Characteristic | Analysis Sample<br>(n=1,013) |  | All available patients<br>(n=2,422) |  |
| --- | --- | --- | --- | --- |
|  | n | (%) | n | (%) |
| Sex at birth |  |  |  |  |
| - Male | 630 | (62%) | 1,490 | (62%) |
| - Female | 383 | (38%) | 931 | (38%) |
| - Missing |  |  | 1 | (0%) |
| Ethnicity |  |  |  |  |
| - White | 795 | (79%) | 1815 | (75%) |
| - South Asian | 87 | (9%) | 273 | (11%) |
| - Black | 73 | (7%) | 170 | (7%) |
| - Mixed | 22 | (2%) | 53 | (2%) |
| - Other | 36 | (4%) | 111 | (4%) |
| - Missing |  |  | 14 | (1%) |
| WHO respiratory support class |  |  |  |  |
| - 4 | 167 | (17%) | 407 | (17%) |
| - 5 | 429 | (42%) | 1,045 | (43%) |
| - 6 | 219 | (22%) | 563 | (23%) |
| - 7-9 | 198 | (20%) | 407 | (17%) |
| Index of Multiple Deprivation quintile |  |  |  |  |
| - 1 (most deprived) | 216 | (21%) | 530 | (22%) |
| - 2 | 217 | (21%) | 566 | (23%) |
| - 3 | 188 | (19%) | 405 | (17%) |
| - 4 | 183 | (18%) | 441 | (18%) |
| - 5 (least deprived) | 209 | (21%) | 469 | (19%) |
| - Missing |  |  | 11 | (0%) |
| Age at admission (years) |  |  |  |  |
| - <30 | 15 | (2%) | 56 | (2%) |
| - 30–39 | 55 | (5%) | 146 | (6%) |
| - 40–49 | 126 | (12%) | 371 | (15%) |
| - 50–59 | 293 | (29%) | 700 | (29%) |
| - 60–69 | 330 | (33%) | 694 | (29%) |
| - 70–79 | 160 | (16%) | 375 | (15%) |
| - 80+ | 34 | (3%) | 80 | (3%) |
| BMI |  |  |  |  |
| - <30kg/m<sup>2</sup> | 423 | (42%) | 725 | (30%) |
| - ≥ 30kg/m<sup>2</sup> | 590 | (58%) | 976 | (40%) |
| - Missing |  |  | 721 | (30%) |
| Presence of baseline comorbidity |  |  |  |  |
| - Cardiac | 467 | (46%) | 1,112 | (46%) |
| - Respiratory | 281 | (28%) | 653 | (27%) |
| - Gastrointestinal | 138 | (14%) | 330 | (14%) |
| - Neurological and psychiatric | 196 | (19%) | 504 | (21%) |
| - Rheumatological | 120 | (12%) | 130 | (5%) |
| - Metabolic/Endocrine/Renal | 119 | (12%) | 294 | (12%) |
| - Malignancy/Haematological | 63 | (6%) | 263 | (11%) |
| EQ5D prior to infection (recall)** | 0.812 | (0.22) | 0.815 | (0.231) |
| - Missing |  |  | 304 | (13%) |
| Hospital site categorisation |  |  |  |  |
| - Assessment | 898 | (89%) | 2,107 | (87%) |
| - Rehabilitation services | 439 | (43%) | 1,246 | (51%) |
| - Mental health services | 357 | (35%) | 980 | (40%) |
| - All patients offered service | 591 | (58%) | 1,577 | (65%) |

\*Analysis Sample is all those with complete data available; \*\*continuous variable: mean (standard deviation.)

Table 1 also provides a summary of the health care pathway variables. Most patients were discharged from a hospital site with comprehensive assessment services (89%), but there was less availability of comprehensive interventions as only 43% of patients were discharged from a site offering a comprehensive rehabilitation service and 35% from a site offering a comprehensive mental health service. In total, 58% of patients were discharged from a site where follow-up services were available to all suitable patients, rather than restricted to a pre-specified sub-group of patients.

In total, 41% (415/1013) of patients had at least one newly diagnosed condition (NDC) at 12-months that was not recorded at baseline hospital admission (Table 2). The number and percentage of the included participants with a NDC of different types at one-year post-hospital admission are also shown in Table 2.

**Table 2.** Newly Diagnosed Conditions (NDCs) at 12 months from discharge.

| Chronic condition | Classification | N (%)<br>1013/100% |
| --- | --- | --- |
| Chronic kidney disease | eGFR < 60 in patient without a previous diagnosis of chronic kidney disease | 94 (9.3%) |
| Diabetes | HbA1c ≥ 6.0% in patient without a previous diagnosis of diabetes | 107 (10.6%) |
| Depression or anxiety | PHQ-9 ≥ 10 or GAD7 > 8 in patient without previous diagnosis of depression or anxiety | 203 (20.0%) |
| Cognitive impairment | MoCA < 23 in patient without previous diagnosis of dementia | 79 (7.8%) |
| Cardiac dysfunction | pBNP ≥ 400 or BNP ≥ 100 in patient without previous diagnosis of heart failure | 41 (4.0%) |
| Total | Any NDC | 415 (41%) |

#### EQ5D-3L Utility Scores

The median EQ5D-3L utility score pre-covid and at the first and second research visits was 0.889 (IQR: 0.744–0.987), 0.753 (0.620–0.891) and 0.752 (0.581–0.893). The median difference in scores between the first research visit and pre-covid was -0.072 (IQR: -0.223– 0.000) and between the second research visit and pre-covid was -0.081 (IQR: -0.232–0.000).

The EQ5D utility score for participants that reported feeling fully recovered from their initial infection with COVID-19 and without an NDC was estimated to be 0.89 (Table 3), somewhat higher than would be expected based on national EQ5D norm data (McNamara et al, 2023). For subjects not feeling fully recovered but without an NDC, their utility was 0.13 units lower at 0.76 (Table 3). The lowest utility score, at 0.66, were for those individuals both not feeling fully recovered and having an NDC. In general, unadjusted scores showed lower utility values for remaining symptoms and NDCs reflecting the association between those health states and higher levels of comorbidity at baseline.

**Table 3.** EQ5D Utility scores by patient perceived recovery and newly diagnosed conditions.

| Health State | Unadjusted mean (SD/n) | Adjusted mean (SE) |
| --- | --- | --- |
| Feel fully recovered with no NDC | 0.87 (0.18 / 339) | 0.89 (0.007) |
| Feel fully recovered with NDC | 0.78 (0.21 / 139) | 0.84 (0.011) |
| Not recovered no NDC | 0.69 (0.25 / 592) | 0.76 (0.011) |
| Not recovered with NDC | 0.57 (0.28 / 475) | 0.66 (0.016) |

\*Unadjusted scores based on observations, adjusted scores based on regression analyses holding all other covariates at mean values

Results for the gamma distributed log-link GLM for EQ5D is presented the Table S2 of Appendix C in the supplementary materials. Clinical and demographic characteristics associated with worse HRQoL were: being female; receiving WHO class 7–9 (includes invasive mechanical ventilation) at hospitalisation compared with class 4 (no supplemental oxygen or other respiratory support) (30); having a respiratory, neurological and/or psychiatric or a rheumatological comorbidity at baseline; and being obese. Conversely, characteristics associated with better HRQoL at one year were a higher pre-COVID utility index summary score and belonging to the least deprived IMD quintile 5 compared to quintile 1.

Controlling for all other covariates, access to comprehensive assessment and comprehensive rehabilitation services were both associated with significantly better HRQoL which results in a significant estimate of quality-of-life benefit for four of the health care pathways estimated (Figure 2).

**Figure 1.**
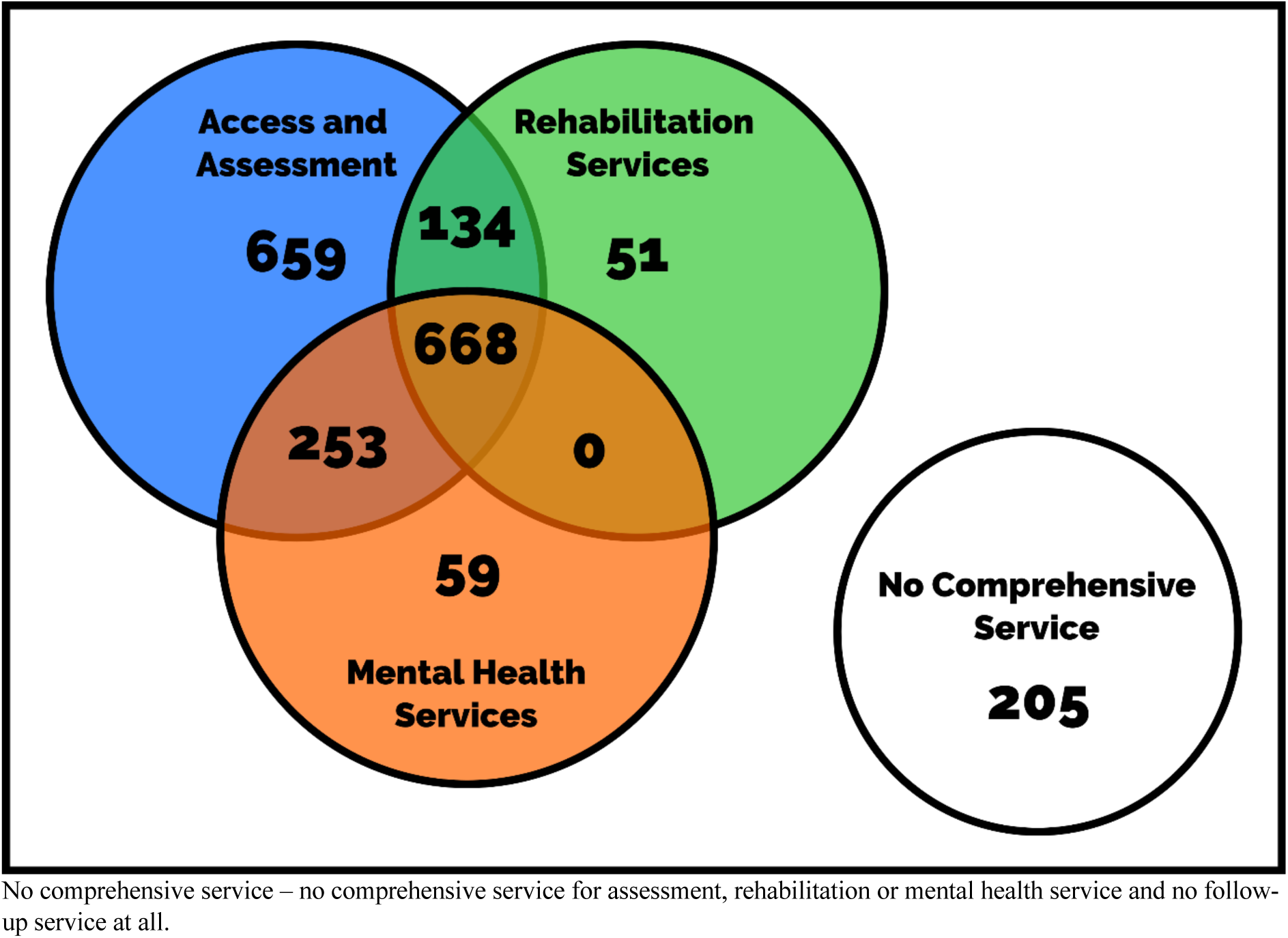
A Euler diagram to highlight patient numbers with access to comprehensive follow-up services for COVID-19 across the metrics of assessment, rehabilitation, and mental health services.

**Figure 2.**
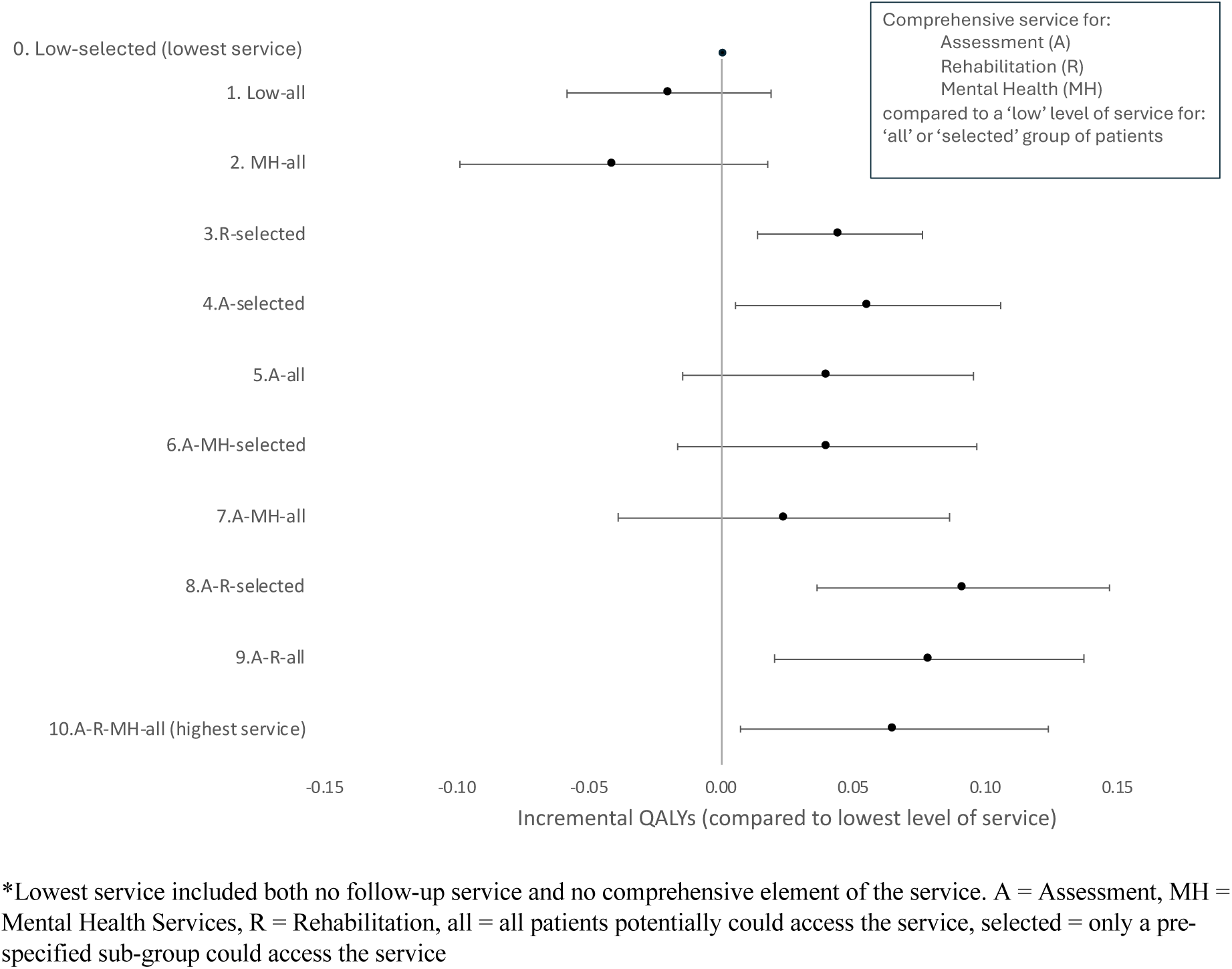
Forest plot of the impact on QALYs of each health care pathway represented in the PHOSP-COVID cohort compared to the lowest level of service available*.

#### Healthcare Resource Use and Associated Costs

The distribution of healthcare resource use costs was right skewed with a mean cost of just over £1,000 per person and values ranging from around £0–£55,000 per person. A gamma distributed log-link GLM for healthcare cost at 12-month is presented in Table S3 of the supplementary materials. Clinical and demographic characteristics that significantly increased healthcare cost were: being female; receiving class 7–9 respiratory support at hospitalisation as opposed to class 4; and having a respiratory or malignancy/ haematological comorbidity at baseline. Conversely, characteristics that significantly reduced healthcare costs were: belonging to IMD quintiles 3–5 compared to the most deprived quintile (quintile 1) and being in age category 30-39 or category 60-69 compared to 50-59. Controlling for all other covariates, none of the healthcare pathway variables had a significant impact on healthcare costs at 12-months post discharge. Figure 3 shows the incremental health service resource costs estimated from the statistical models for each of ten pathways compared to the lowest service pathway as a forest plot.

**Figure 3.**
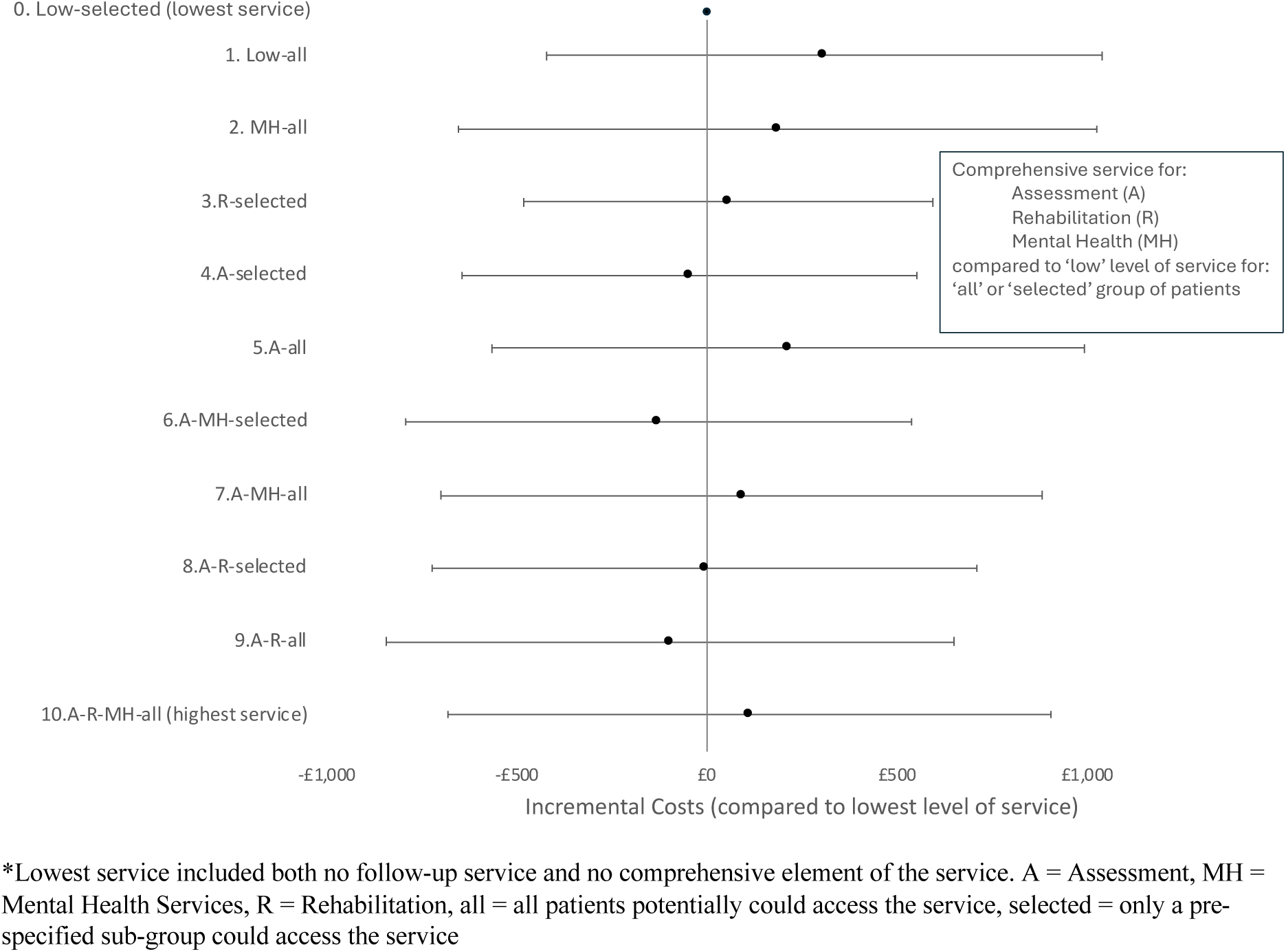
Forest plot of the impact of each health care pathway in PHOSP-COVID on Cost compared to the lowest service offering.

### Cost-effectiveness of healthcare pathways

The estimated costs and effects based on the statistical models from Tables S2 & S3 of Appendix C are presented in Table 4 for each of the healthcare pathways represented in PHOSP-COVID. The highest healthcare pathway has an estimated incremental cost-effectiveness ratio of £1,700 per QALY with confidence interval in the dominant quadrant of the plane up to £24,800 per QALY (Figure 4).

**Figure 4.**
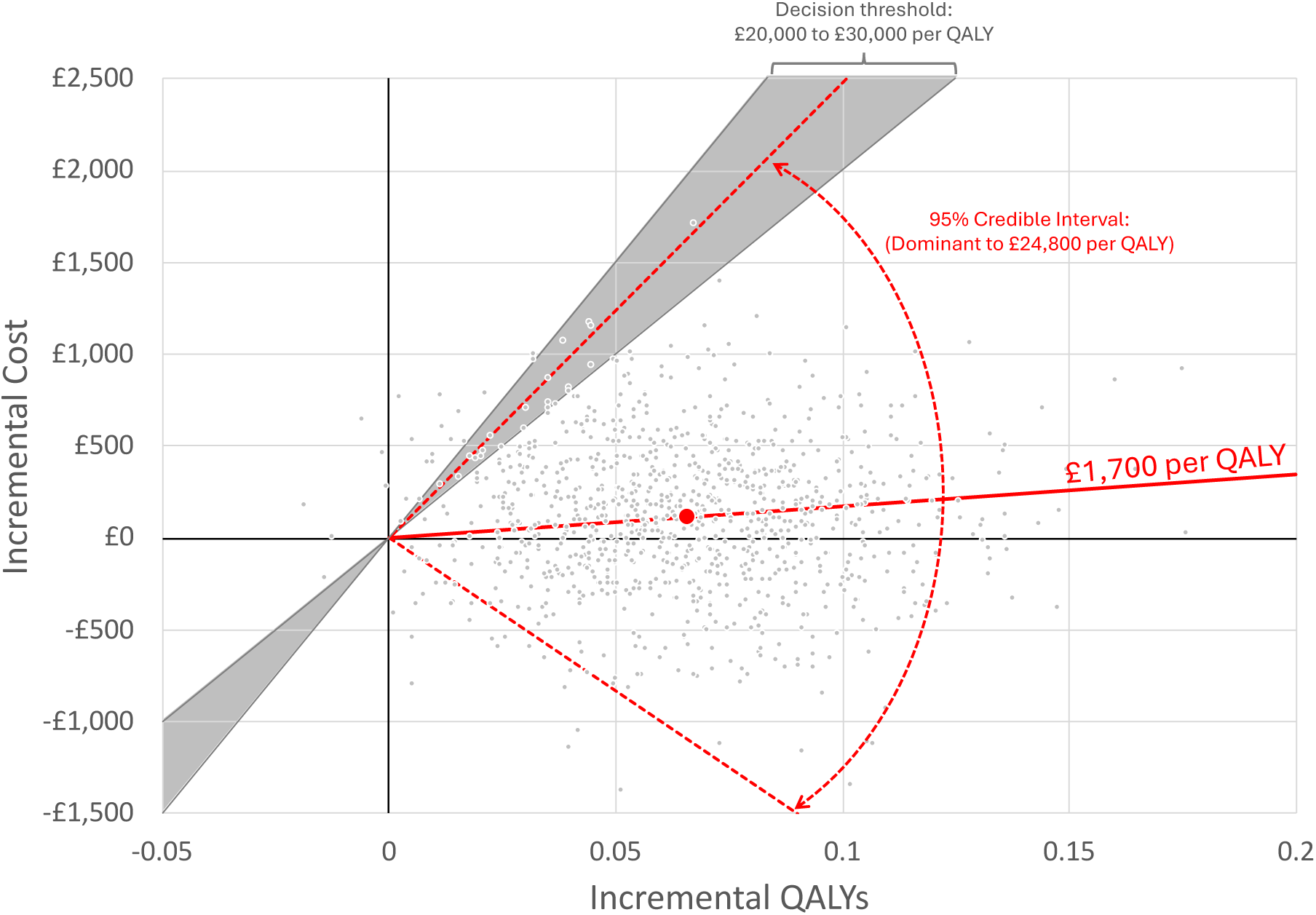
Comparison of the highest service pathway (10) to the lowest service pathway (0) on the cost-effectiveness plane. Red circle shows the point estimate of cost-effectiveness, slope of red solid line shows incremental cost-effectiveness ratio, grey dots show uncertainty and red dotted lines show the 95% credible interval for cost-effectiveness.

**Table 4.** Estimated QALYs at one year and costs for each of the unique 11 health care pathways available at PHOSP-COVID sites. The presence of each of the four main effects (assessment, rehabilitation, mental health, service offered to all) are shown in the first four columns. The last column of the table shows the label for that pathway

| Comprehensive services |  |  | All patients invited to attend clinic?<br>No (selected)<br>Yes (all) | QALY | SE | Cost (£) | SE | label |
| --- | --- | --- | --- | --- | --- | --- | --- | --- |
| Assessment (A) | Rehabilitation (R) | Mental health (MH) |  |  |  |  |  |  |
| No | No | No | No | 0.725 | 0.026 | 755 | 276 | 0. Low-selected |
| No | No | No | Yes | 0.706 | 0.032 | 1032 | 479 | 1. Low-all |
| No | No | Yes | Yes | 0.686 | 0.038 | 914 | 484 | 2. MH-all |
| No | Yes | No | No | 0.770 | 0.026 | 782 | 392 | 3. R-selected |
| Yes | No | No | No | 0.780 | 0.013 | 720 | 237 | 4. A-selected |
| Yes | No | No | Yes | 0.764 | 0.014 | 983 | 305 | 5. A-all |
| Yes | No | Yes | No | 0.765 | 0.016 | 637 | 252 | 6. A-MH-selected |
| Yes | No | Yes | Yes | 0.748 | 0.018 | 870 | 286 | 7. A-MH-all |
| Yes | Yes | No | No | 0.816 | 0.015 | 744 | 300 | 8. A-R-selected |
| Yes | Yes | No | Yes | 0.803 | 0.016 | 659 | 297 | 9. A-R-all |
| Yes | Yes | Yes | Yes | 0.789 | 0.012 | 900 | 269 | 10. A-R-MH-all |

#### Sensitivity analysis weights

The regression models of Tables S2 & S3 in Appendix C of the supplementary materials were re-estimated using propensity score weights to adjust for potential missing at random effects of the missing data and to adjust the PHOSP cohort to look more representative of the true hospital population using ISARIC (26) as a reference population. Tables S4 & S5 in Appendix C of the supplementary materials show the re-weighted regression analyses.

Overall, coefficients from the regression models were not substantially different when using the weighted analyses, consequently neither was the estimated cost-effectiveness of the highest level of service (which reduced slightly to under £1,000 per QALY). Alternative specifications of the models, including using non recovery and existence of NDCs as mediating variables and as control variables, did not change the results markedly (results not shown).

## Discussion

We report for the first time that healthcare pathways for adult survivors of a hospital admission for COVID-19 appear to be clinically effective if they offer a comprehensive service. The most comprehensive services, where all patients could potentially access comprehensive assessment, rehabilitation, and mental health services, were clinically effective when compared with either no service or ‘light touch’ (lowest) services (Figure 2). The most comprehensive service was also cost effective compared to no service or ’light touch’ services (lowest) (Figure 4) with an estimated cost per QALY of £1,700 (95% uncertainty interval: dominated to £24,800).

Our results particularly support the effectiveness of a comprehensive assessment and availability of multi-dimensional rehabilitation. To our knowledge this is the first description highlighting clinical effectiveness of a complex/ comprehensive assessment. The comprehensive assessment included a face-to-face option, a multi-system approach, complex diagnostics, and availability of a multi-disciplinary and inter-speciality team meeting. To date, the type of follow-up assessment provided (for any of the healthcare pathways) has been based on expert opinion. The NHS England Long Covid service specification is one of very few internationally to be designed and implemented at a national level (31). Our data support the components described in the service specification.

Our data suggest that multi-dimensional rehabilitation is clinically effective, which supports systematic reviews of small-scale randomised controlled trials investigating rehabilitation versus usual care that suggest effectiveness (14), although definitive trials are needed. Our data are perhaps less certain for mental health interventions in isolation and most exercise-based rehabilitation programmes also contain interventions to support mental health and some are integrated which we may not have captured from the survey. A systematic review of registered trials for interventions for mental health, cognition, and psychological wellbeing in Long Covid (32) highlighted that the breadth and scope of research remains limited. Our data highlight a significant new burden of symptoms suggestive of anxiety and depression (the challenges of interpretation of the questionnaires in a physically unwell population notwithstanding) and therefore highlight the urgent need for interventions to improve both physical and mental health. The categorisation of services offered to all patients or a select group of patients did not seem to have a large impact on the results in our cohort study, but we would recommend all patients with potential need to have access to services rather than a pre-specified criterion.

Although our data show clinical effectiveness for the more comprehensive services, there is a balance between the cost of a comprehensive service for all patients to access versus either limiting to those with the most severe acute disease or only providing a light touch service such as a one-off telephone call or no service at all. We report positive data on cost per QALY suggesting that the most comprehensive service is both clinically and cost-effective based on commonly accepted thresholds for cost-effectiveness in the £20-30,000 per QALY range (33).

In all, only 29% of patients report feeling fully recovered from COVID at one year after discharge. NDCs were apparent in 46% of participants which could account for remaining symptoms which leaves 39% reporting sustained symptoms at one-year with no clear cause – this is the closest group in our data to the definition of Post-Covid-19 condition (Long Covid) by WHO (34). However, it is an underestimate of the prevalence of Long Covid as it assumes that, in patients with a newly diagnosed condition, these conditions fully accounts for their persistent symptoms which is unlikely. In addition to our previous reports of low rates of patient-perceived recovery at one year after discharge from hospital in the PHOSP-COVID study (35), we highlight a large new health burden of NDCs such as diabetes, new mental health symptoms, and cognitive impairment. While we concede that some of these could have been pre-existing before COVID-19 but undiagnosed, many will be as a result of (or exacerbated by) COVID-19. These long-term consequences of COVID-19 require optimised treatment, which supports the need for multi-speciality expertise being available for Long Covid clinics (2).

### Strength and Limitations

The strengths of the data are the detailed objective follow-up of a large number of participants alongside detailed characterisation of the Long Covid follow up at their hospital site. Selection and survivor bias (cohort are survivors to one year after discharge) were mitigated by modelling to both the larger PHOSP-COVID cohort, and to the ISARIC data set (a larger cohort of patients admitted into a UK hospital for COVID-19).

However, there are important limitations to be considered. Although attempts have been made to control for observed confounding, given this is an observational study, it is likely that unobserved confounding remains. There was difficulty in determining the precise level of post-COVID-19 services on offer for individual participants as this information was mapped at the site level from survey data, and is therefore not a direct assessment of services. The participants were discharged from hospital between February 2020 to 31^st^ March 2021 and were therefore mostly unvaccinated prior to hospital admission and before use of most therapeutics for acute COVID-19. The hospital admission data was not retrieved from national NHS linkage but was retrieved by researchers from the patients’ medical records. For example, an admission at a different location may not have been known about if the participant did not recall it. Our data is for patients with severe COVID-19 and cannot be directly extrapolated to non-hospitalised cases of Long Covid. However, the comprehensive clinical care model is applicable as described by the NHS-England service specification.

Clinical and cost-effectiveness requires further evaluation in the non-hospitalised population. Despite higher vaccination rates, better acute treatments for COVID-19 and new variants of the disease, our data likely remains applicable for contemporary patients who are nevertheless serious enough to be hospitalised for their acute infection and some groups of patients such as the immunocompromised population have remained at the same high risk of severe disease through the pandemic (36).

### Clinical implications

To date Long Covid care is heterogenous across the UK and Internationally. Our data supports the need for proactive care, and for a clinically and cost-effective comprehensive care model for assessment, rehabilitation and mental health services. This is predominantly to improve health-related quality of life for individuals which is similarly reduced in our data compared to other long-term conditions (37). However, there are additional benefits to dedicated Long Covid clinics such as developing teams of healthcare professionals that are experts in this complex multi-system disease and who could collectively run clinical trials of much-needed treatments in eligible patients. Other benefits include establishing correct coding of health records and helping industry understand the healthcare models their products would be prescribed within if clinical trials were successful.

## Summary

In summary, comprehensive healthcare models for assessment, rehabilitation and mental health services for adult survivors of a hospital admission for COVID-19 are estimated to be clinically effective and cost-effective compared to commonly accepted thresholds. Further work needs to be extended to healthcare models for the larger group of non-hospitalised patients who develop Long Covid.

## Data Availability

The PHOSP-COVID study website (https://www.phosp.org) contains an overview of the study, resources, information about people involved, and publications. Research activity using the study is organised across a series of Working Groups (Figure 3). These were established at the outset of the study to coordinate research, minimise duplication of efforts, and facilitate communication across research and clinical specialties. Researchers interested in undertaking research using PHOSP-COVID are encouraged to contact the relevant Working Group leads (https://www.phosp.org/working-group/) in the first instance. The data are currently held in the Outbreak Data Analysis Platform (ODAP, https://odap.ac.uk/). Researchers seeking to access these data are directed to https://www.phosp.org/resource/ for information and forms. Correspondence to be directed to Dr Rachael A Evans, the Co-Principal Investigator of PHOSP-COVID study phosp{at}leicester.ac.uk.

## Acknowledgments

This study would not have been possible without the existence of the PHOSP dataset and all the participants who have given their time and support. We thank all the participants and their families. We thank the many research administrators, health-care and social-care professionals who contributed to setting up and delivering the study at all of the NHS trusts/Health boards and research institutions across the UK, as well as all the supporting staff at the NIHR Clinical Research Network, Health Research Authority, Research Ethics Committee, Department of Health and Social Care, Public Health Scotland, and Public Health England, and support from the ISARIC Coronavirus Clinical Characterisation Consortium. We thank Kate Holmes at the NIHR Office for Clinical Research Infrastructure (NOCRI) for her support in coordinating the charities group. The PHOSP-COVID industry framework was formed to provide advice and support in commercial discussions, and we thank the Association of the British Pharmaceutical Industry as well NOCRI for coordinating this. We are very grateful to all the charities that have provided insight to the study: Action Pulmonary Fibrosis, Alzheimer’s Research UK, Asthma + Lung UK, British Heart Foundation, Diabetes UK, Cystic Fibrosis Trust, Kidney Research UK, MQ Mental Health, Muscular Dystrophy UK, Stroke Association Blood Cancer UK, McPin Foundations, and Versus Arthritis. We thank the NIHR Leicester Biomedical Research Centre patient and public involvement group and Long Covid Support.

