## Supplementary materials for "Clinical and cost-effectiveness of diverse post-hospitalisation pathways for COVID-19: A UK evaluation utilising the PHOSP-COVID cohort"

#### Appendix A: PHOSP-COVID Collaborative Group

### PHOSP-COVID Collaborative Group

**Core Management Group**

*Chief Investigator* C E Brightling, *Members* R A Evans (Lead Co-I), L V Wain (Lead Co-I), J D Chalmers, V C Harris, L P Ho, A Horsley, M Marks, K Poinasamy, B Raman, A Shikotra, A Singapuri

**PHOSP-COVID Study Central Coordinating Team**

C E Brightling (Chief Investigator), R A Evans (*Lead Co-I*), L V Wain (*Lead Co-I*), R Dowling, C Edwardson, O Elneima, S Finney, N J Greening, B Hargadon, V C Harris, L Houchen--Wolloff, O C Leavy, H J C McAuley, C Overton, T Plekhanova, R M Saunders, M Sereno, A Singapuri, A Shikotra, C Taylor, S Terry, C Tong, B Zhao

**Steering Committee**

*Co-chairs* D Lomas, E Sapey*, Institution representatives* C Berry, C E Bolton, N Brunskill, E R Chilvers, R Djukanovic, Y Ellis, D Forton, N French, J George, N A Hanley, N Hart, L McGarvey, N Maskell, H McShane, M Parkes, D Peckham, P Pfeffer, A Sayer, A Sheikh, A A R Thompson, N Williams and core management group representation

**Executive Board**

*Chair* C E Brightling, representation from the core management group, each working group and platforms

#### Platforms

**Bioresource**

W Greenhalf (*Co-Lead*), M G Semple (*Co-Lead*), M Ashworth, H E Hardwick, L Lavelle-Langham, W Reynolds, M Sereno, R M Saunders, A Singapuri, V Shaw, A Shikotra, B Venson, L V Wain

**Data Hub**

A B Docherty (*Co-Lead*), E M Harrison (*Co-Lead*), A Sheikh (*Co-Lead*), J K Baillie, C E Brightling, L Daines, R Free, R A Evans, S Kerr, O C Leavy, N I Lone, D Lozano-Rojas, H J C McAuley, K Ntotsis, R Pius, J Quint, M Richardson, , M Sereno, M Thorpe, L V Wain

**Genetic Analysis**

L V Wain (Co-Lead), J K Baillie (*Co-Lead*), N Avramidis, E Coughlan, B Guillen-Guio, O C Leavy, E Pairo-Castineira, K Rawlik

**Imaging Alliance**

M Halling-Brown (*Co-Lead*), F Gleeson (*Co-Lead*), J Jacob (*Co-Lead*), S Neubauer (*Co-Lead*) B Raman (*Co-Lead*) S Siddiqui (*Co-Lead*) J M Wild (*Co-Lead*), S Aslani, G Baxter, M Beggs, C Bloomfield, M P Cassar, A Chiribiri, E Cox, D J Cuthbertson, M Halling-Brown, V M Ferreira, L Finnigan, S Francis, P Jezzard, G J Kemp, H Lamlum, E Lukaschuk, C Manisty, G P McCann , C McCracken, K McGlynn , R Menke , C A Miller , A J Moss, T E Nichols, C Nikolaidou , C O'Brien , G Ogbole, B Rangelov, D P O'Regan , A Pakzad, S Piechnik , S Plein, I Propescu, A A Samat, L Saunders, Z B Sanders, R Steeds, T Treibel, E M Tunnicliffe, M Webster, J Willoughby, J Weir McCall, C Xie, M Xu

**Omics**

L V Wain (*Co-Lead)*, J K Baillie (*Co-Lead*), H Baxendale, C E Brightling, M Brown, J D Chalmers, R A Evans, B Gooptu, W Greenhalf, H E Hardwick, R G Jenkins, D Jones, I Koychev, C Langenberg, A Lawrie, P L Molyneaux, A Shikotra, J Pearl, M Ralser, N Sattar, R M Saunders, J T Scott, T Shaw, D Thomas, D Wilkinson

#### Working Groups

**Airways**

L G Heaney (*Co-Lead*), A De Soyza (*Co-Lead*), D Adeloye, C E Brightling, J S Brown, J Busby, J D Chalmers, C Echevarria, L Daines, O Elneima, RA Evans, J Hurst, P Novotny, C Nicolaou, P Pfeffer, K Poinasamy, J Quint, I Rudan, E Sapey, M Shankar-Hari, A Sheikh, S Siddiqui, S Walker, B Zheng

**Brain**

J R Geddes (*Lead*), M Hotopf *(Co-Lead),* K Abel, R Ahmed, L Allan, C Armour, D Baguley, D Baldwin, C Ballard, K Bhui, G Breen, K Breeze, M Broome, T Brugha, E Bullmore, D Burn, F Callard, J Cavanagh, T Chalder, D Clark, A David, B Deakin, H Dobson, B Elliott, J Evans, RA Evans, R Francis, E Guthrie, P Harrison, M Henderson, A Hosseini, N Huneke, M Husain, T Jackson, I Jones, T Kabir, P Kitterick, A Korszun, I Koychev, J Kwan, A Lingford-Hughes, P Mansoori, H McAllister-Williams, K McIvor, B Michael, L Milligan, R Morriss, E Mukaetova-Ladinska, K Munro, A Nevado-Holgado, T Nicholson, C Nicolaou, S Paddick, C Pariante, J Pimm, K Saunders, M Sharpe, G Simons, J P Taylor, R Upthegrove, S Wessely

**Cardiac**

G P McCann (*Lead*), S Amoils, C Antoniades, A Banerjee, A Bularga, C Berry, P Chowienczyk, J P Greenwood, A D Hughes, K Khunti, C Lawson, N L Mills, A J Moss, S Neubauer, B Raman, A N Sattar, C L Sudlow, M Toshner,

**Immunology**

P J M Openshaw (*Lead*), D Altmann, J K Baillie, R Batterham, H Baxendale, N Bishop, C E Brightling, P C Calder, C M Efstathiou, R A Evans, J L Heeney, T Hussell, P Klenerman, F Liew, J M Lord, P Moss, S L Rowland-Jones, W Schwaeble, M G Semple, R S Thwaites, L Turtle, L V Wain, S Walmsley, D Wraith

**Intensive Care**

M J Rowland (*Lead*), A Rostron (*Co-Lead*), J K Baillie, B Connolly, A B Docherty, N I Lone, D F McAuley, D Parekh, A Rostron, J Simpson, C Summers

**Lung Fibrosis**

R G Jenkins (*Co-Lead*), J Porter (*Co-Lead*), R J Allen, R Aul, J K Baillie, S Barratt, P Beirne, J Blaikley, R C Chambers, N Chaudhuri, C Coleman, E Denneny, L Fabbri, P M George, M Gibbons, F Gleeson, B Gooptu, B Guillen-Guio, I Hall, N A Hanley, L P Ho, E Hufton, J Jacob, I Jarrold, G Jenkins, S Johnson, M G Jones, S Jones, F Khan, P Mehta, J Mitchell, P L Molyneaux, J E Pearl, K Piper Hanley, K Poinasamy, J Quint, D Parekh, P Rivera-Ortega, L C Saunders, M G Semple, J Simpson, D Smith, M Spears, L G Spencer, S Stanel, I Stewart, A A R Thompson, D Thickett, R Thwaites, L V Wain, S Walker, S Walsh, J M Wild, D G Wootton, L Wright

**Metabolic**

S Heller (*Co-Lead*), M J Davies (*Co-Lead*), H Atkins, S Bain, J Dennis, K Ismail, D Johnston, P Kar, K Khunti, C Langenberg, P McArdle, A McGovern, T Peto, J Petrie, E Robertson, N Sattar, K Shah, J Valabhji, B Young

**Pulmonary and Systematic Vasculature**

L S Howard (*Co-Lead*), Mark Toshner (*Co-Lead*), C Berry, P Chowienczyk, A Lawrie, O C Leavy, J Mitchell, J Newman, L Price, J Quint, A Reddy, J Rossdale, N Sattar, C Sudlow, A A R Thompson, J M Wild, M Wilkins

**Rehabilitation, Sarcopenia and Fatigue**

S J Singh (*Co-Lead*), W D-C Man (*Co-Lead*), J M Lord (*Co-Lead*), N J Greening (*Co-Lead*), T Chalder (*Co-Lead*), J T Scott (*Co-Lead*), N Armstrong, E Baldry, M Baldwin, N Basu, M Beadsworth, L Bishop, C E Bolton, A Briggs, M Buch, G Carson, J Cavanagh, H Chinoy, C Dawson, E Daynes, S Defres, R A Evans, L Gardiner, P Greenhaff, S Greenwood, M Harvie, L HOuchen-Wolloff, M Husain, S MacDonald, A McArdle, H J C McAuley, A McMahon, M McNarry, G Mills, C Nolan, K O'Donnell, D Parekh, Pimm, J Sargent, L Sigfrid, M Steiner, D Stensel, A L Tan, I Vogiatzis, J Whitney, D Wilkinson, D Wilson, M Witham, D G Wootton, T Yates

**Renal**

D Thomas (*Lead*), N Brunskill (*Co-Lead*), S Francis (*Co-Lead*), S Greenwood (*Co-Lead*), C Laing (*Co-Lead*), K Bramham, P Chowdhury, A Frankel, L Lightstone, S McAdoo, K McCafferty, M Ostermann, N Selby, C Sharpe, M Willicombe

**Patient Public Engagement Group**

L Houchen-Wolloff (*Lead*), J Bunker, R Gill, C Hastie, R Nathu, N Rogers, N Smith

#### Local Clinical Centre PHOSP-COVID trial staff

(listed in alphabetical order)

**Airedale NHS Foundation Trust**

A Shaw (PI), L Armstrong, B Hairsine, H Henson, C Kurasz, L Shenton

**Aneurin Bevan University Health Board**

S Fairbairn (PI), A Dell, N Hawkings, J Haworth, M Hoare, A Lucey, V Lewis, G Mallison, H Nassa, C Pennington, A Price, C Price, A Storrie, G Willis, S Young

**Barts Health NHS Trust & Queen Mary University of London**

P Pfeffer (PI), K Chong-James, C David, W Y James, C Manisty, A Martineau, O Zongo

**Barnsley Hospital NHS Foundation Trust**
A Sanderson (PI)

**Belfast Health and Social Care Trust & Queen's University Belfast**
L G Heaney (PI), C Armour, V Brown, T Craig, S Drain, B King, N Magee, D McAulay, E Major, L McGarvey, J McGinness, R Stone

**Betsi Cadwaladr University Health Board**
A Haggar (PI), A Bolger, F Davies, J Lewis, A Lloyd, R Manley, E McIvor, D Menzies, K Roberts, W Saxon, D Southern, C Subbe, V Whitehead

**Borders General Hospital, NHS Borders**

H El-Taweel (PI), J Dawson, L Robinson

**Bradford Teaching Hospitals NHS Foundation Trust**

D Saralaya (PI), L Brear, K Regan, K Storton

**Cambridge University Hospitals NHS Foundation Trust, NIHR Cambridge Clinical Research Facility & University of Cambridge**
J Fuld (PI), A Bermperi, I Cruz, K Dempsey, A Elmer, H Jones, S Jose, S Marciniak, M Parkes, C Ribeiro, J Taylor, M Toshner, L Watson, J Weir McCall, J Worsley

**Cardiff and Vale University Health Board**

R Sabit (PI), L Broad, A Buttress, T Evans, M Haynes, L Jones, L Knibbs, A McQueen, C Oliver, K Paradowski, J Williams

**Chesterfield Royal Hospital NHS Trust**

E Harris (PI), C Sampson

**Cwm Taf Morgannwg University Health Board**

C Lynch (PI), E Davies, C Evenden , A Hancock, K Hancock, M Rees , L Roche, N Stroud, T Thomas-Woods

**East Cheshire NHS Trust**

M Babores (PI), J Bradley-Potts, M Holland, N Keenan, S Shashaa, H Wassall

**East Kent Hospitals University NHS Foundation Trust**

E Beranova (PI), H Weston (PI), T Cosier, L Austin, J Deery, T Hazelton, C Price, H Ramos, R Solly, S Turney

**Gateshead NHS Trust**

L Pearce (PI), W McCormick, S Pugmire, W Stoker, A Wilson

**Guy's and St Thomas' NHS Foundation Trust**
N Hart (PI), LA Aguilar Jimenez, G Arbane, S Betts, K Bisnauthsing, A Dewar, P Chowdhury, A Chiribiri, A Dewar, G Kaltsakas, H Kerslake, MM Magtoto, P Marino, LM Martinez, C O'Brien, M Ostermann, J Rossdale, TS Solano, E Wynn

**Hampshire Hospitals NHS Foundation Trust**
N Williams (PI), W Storrar (PI), M Alvarez Corral, A Arias, E Bevan, D Griffin, J Martin, J Owen, S Payne, A Prabhu, A Reed, C Wrey Brown

**Harrogate and District NHD Foundation Trust**

C Lawson (PI), T Burdett, J Featherstone, A Layton, C Mills, L Stephenson,

**Hull University Teaching Hospitals NHS Trust & University of Hull**
N Easom (PI), P Atkin, K Brindle, M G Crooks, K Drury, R Flockton, L Holdsworth, A Richards, D L Sykes, S Thackray-Nocera, C Wright

**Hywel Dda University Health Board**

K E Lewis (PI), A Mohamed (PI), G Ross (PI), S Coetzee, K Davies, R Hughes, R Loosley, L O'Brien, Z Omar, H McGuinness, E Perkins, J Phipps, A Taylor, H Tench, R Wolf-Roberts

**Imperial College Healthcare NHS Trust & Imperial College London**
L S Howard (PI), O Kon (PI), D C Thomas (PI), S Anifowose, L Burden, E Calvelo, B Card, C Carr, E R Chilvers, D Copeland, P Cullinan, P Daly, C M Efstathiou, L Evison, T Fayzan, H Gordon, S Haq, R G Jenkins, C King, F Liew, K March, M Mariveles, L McLeavey, N Mohamed, S Moriera, U Munawar, J Nunag, U Nwanguma, L Orriss- Dib, D P O'Regan, A Ross, M Roy, E Russell, K Samuel, J Schronce, N Simpson, L Tarusan, C Wood, N Yasmin

**Kettering General Hospital NHS Trust**

R Reddy (PI), A-M, Guerdette, M Hewitt, K Warwick, S White

**King's College Hospital NHS Foundation Trust & Kings College London**
A M Shah (PI), C J Jolley (PI), O Adeyemi, R Adrego, H Assefa-Kebede, J Breeze, M Brown, S Byrne, T Chalder, A Chiribiri, P Dulawan, N Hart, A Hayday, A Hoare, A Knighton, M Malim, C O'Brien, S Patale, I Peralta, N Powell, A Ramos, K Shevket, F Speranza, A Te

**Leeds Teaching Hospitals & University of Leeds**
P Beirne (PI), A Ashworth, J Clarke, C Coupland, M Dalton, E Wade, C Favager, J Greenwood, J Glossop, L Hall, T Hardy, A Humphries, J Murira, D Peckham, S Plein, J Rangeley, G Saalmink, A L Tan, B Whittam, N Window, J Woods,

**Lewisham & Greenwich NHS Trust**

G Coakley (PI)

**Liverpool University Hospitals NHS Foundation Trust & University of Liverpool**

D G Wootton (PI), L Turtle (PI), L Allerton, AM All, M Beadsworth, A Berridge, J Brown, S Cooper, A Cross, D J Cuthbertson, S Defres, S L Dobson, J Earley, N French, W Greenhalf, H E Hardwick, K Hainey, J Hawkes, V Highett, S Kaprowska, G J Kemp, AL Key, S Koprowska, L Lavelle-Langham, N Lewis-Burke, G Madzamba, F Malein, S Marsh, C Mears, L Melling, M J Noonan, L Poll, J Pratt, E Richardson, A Rowe, M G Semple, V Shaw, K A Tripp, B Vinson, L O Wajero, S A Williams-Howard, J Wyles

**London North West University Healthcare NHS Trust**

S N Diwanji (PI), P Papineni (PI), S Gurram, S Quaid, G F Tiongson, E Watson

**Manchester University NHS Foundation Trust & University of Manchester**

B Al-Sheklly (PI), A Horsley (PI), C Avram, P Barran, J Blaikely, M Buch, N Choudhury, D Faluyi, T Felton, T Gorsuch, N A Hanley, T Hussell, Z Kausar, C A Miller, N Odell, R Osbourne, K Piper Hanley, K Radhakrishnan, S Stockdale, D Trivedi

**Newcastle upon Tyne Hospitals NHS Foundation Trust & University of Newcastle**

A De Soyza (PI), C Echevarria (PI), A Ayoub, J Brown, G Burns, G Davies, H Fisher, C Francis, A Greenhalgh, P Hogarth, J Hughes, K Jiwa, G Jones, G MacGowan, D Price, A Sayer, J Simpson, H Tedd, S Thomas, S West, M Witham, S Wright, A Young

**NHS Dumfries and Galloway**

M J McMahon (PI), P Neill

**NHS Greater Glasgow and Clyde Health Board & University of Glasgow**

D Anderson (PI), H Bayes (PI), C Berry (PI), D Grieve (PI), I B McInnes (PI), N Basu, A Brown, A Dougherty, K Fallon, L Gilmour, K Mangion, A Morrow, K Scott, R Sykes, R Touyz

**NHS Highland**

E K Sage (PI), F Barrett, A Donaldson

**NHS Lanarkshire**

M Patel (PI), D Bell, A Brown, M Brown, R Hamil, K Leitch, L Macliver, J Quigley, A Smith, B Welsh

**NHS Lothian & University of Edinburgh**

G Choudhury (PI), J K Baillie, S Clohisey, A Deans, A B Docherty, J Furniss, E M Harrison, S Kelly, N I Lone, D E Newby, A Sheikh

**NHS Tayside & University of Dundee**

J D Chalmers (PI), D Connell, A Elliott, C Deas, J George, S Mohammed, J Rowland, A R Solstice, D Sutherland, C J Tee

**North Bristol NHS Trust & University of Bristol**

N Maskell (PI), D Arnold, S Barrett, H Adamali, A Dipper, S Dunn, A Morley, L Morrison, L Stadon, S Waterson, H Welch

**North Middlesex Hospital NHS Trust**

B Jayaraman (PI), T Light

**Nottingham University Hospitals NHS Trust & University of Nottingham**

C E Bolton (PI), P Almeida, J Bonnington, M Chrystal, E Cox, C Dupont, S Francis, P Greenhaff, A Gupta, L Howard, W Jang, S Linford, L Matthews, R Needham, A Nikolaidis, S Prosper, K Shaw, A K Thomas

**Oxford University Hospitals NHS Foundation Trust & University of Oxford**

L P Ho (PI), N M Rahman (PI), M Ainsworth, A Alamoudi, M Beggs, A Bates, A Bloss, A Burns, P Carter, M Cassar, K M Channon, J Chen, F Conneh, T Dong, R I Evans, E Fraser, X Fu, J R Geddes, F Gleeson, P Harrison, M Havinden-Williams, P Jezzard, N Kanellakis, I Koychev, P Kurupati, X Li, E Lukaschuk, K McGlynn, H McShane, C Megson, K Motohashi, S Neubauer, D Nicoll, G Ogg, E Pacpaco, M Pavlides, Y Peng, N Petousi, J Propescu, N Rahman, B Raman, M J Rowland, K Saunders, M Sharpe, N Talbot, E Tunnicliffe

**Royal Brompton and Harefield Clinical Group, Guy's and St Thomas' NHS Foundation Trust.**

W D-C Man (PI), B Patel (PI), R E Barker, D Cristiano, N Dormand, M Gummadi, S Kon, K Liyanage, C M Nolan, S Patel, O Polgar, P Shah, S J Singh, J A Walsh

**Royal Free London NHS Foundation Trust**

J Hurst (PI), H Jarvis (PI), S Mandal (PI), S Ahmad, S Brill, L Lim, D Matila, O Olaosebikan, C Singh

**Royal Papworth Hospital NHS Foundation Trust**

M Toshner (PI), H Baxendale, L Garner, C Johnson, J Mackie, A Michael, J Pack, K Paques, H Parfrey, J Parmar

**Salford Royal NHS Foundation Trust**

N Diar Bakerly (PI), P Dark, D Evans, E Hardy, A Harvey, D Holgate, S Knight, N Mairs, N Majeed, L McMorrow, J Oxton, J Pendlebury, C Summersgill, R Ugwuoke, S Whittaker

**Salisbury NHS Foundation Trust**

W Matimba-Mupaya (PI), S Strong-Sheldrake

**Sheffield Teaching NHS Foundation Trust & University of Sheffield**

S L Rowland-Jones (PI), A A R Thompson (Co PI), J Bagshaw, M Begum, K Birchall, R Butcher, H Carborn, F Chan, K Chapman, Y Cheng, L Chetham, C Clark, Z Coburn, J Cole, M Dixon, A Fairman, J Finnigan, L Finnigan, H Foot, D Foote, A Ford, R Gregory, K Harrington, L Haslam, L Hesselden, J Hockridge, A Holbourn, B Holroyd-Hind, L Holt, A Howell, E Hurditch, F Ilyas, C Jarman, A Lawrie, E Lee, J-H Lee, R Lenagh, A Lye, I Macharia, M Marshall, A Mbuyisa, J McNeill, S Megson, J Meiring, L Milner, S Misra, H Newell, T Newman, C Norman, L Nwafor, D Pattenadk, M Plowright, J Porter, P Ravencroft, C Roddis, J Rodger, P Saunders, J Sidebottom, J

Smith, L Smith, N Steele, G Stephens, R Stimpson, B Thamu, N Tinker, K Turner, H Turton, P Wade, S Walker, J Watson, J M Wild, I Wilson, A Zawia

**St George's University Hospitals NHS Foundation Trust**
R Aul (PI), M Ali, A Dunleavy (PI), D Forton, N Msimanga, M Mencias, T Samakomva, S Siddique, J Teixeira, V Tavoukjian

**Sherwood Forest Hospitals NHS Foundation Trust**
J Hutchinson (PI), L Allsop, K Bennett, P Buckley, M Flynn, M Gill, C Goodwin, M Greatorex, H Gregory, C Heeley, L Holloway, M Holmes, J Kirk, W Lovegrove, TA Sewell, S Shelton, D Sissons, K Slack, S Smith, D Sowter, S Turner, V Whitworth, I Wynter

**Shropshire Community Health NHS Trust**
L Warburton (PI), S Painter, J Tomlinson

**Somerset NHS Foundation Trust**
C Vickers (PI), T Wainwright, D Redwood, J Tilley, S Palmer

**Swansea Bay University Health Board**
G A Davies (PI), L Connor, A Cook, T Rees, F Thaivalappil, C Thomas

**Tameside and Glossop Integrated Care NHS Foundation**

A Butt (PI), M Coulding, H Jones, S Kilroy, J McCormick, J McIntosh, H Savill, V Turner, J Vere

**The Great Western Hospital Foundation Trust**

E Fraile (PI), J Ugoji

**The Hillingdon Hospitals NHS Foundation Trust**
S S Kon (PI), H Lota, G Landers, M Nasseri, S Portukhay

**The Rotherham NHS Foundation Trust**

A Hormis (PI), A Daniels, J Ingham, L Zeidan

**United Lincolnshire Hospitals NHS Trust**
M Chablani (PI), L Osborne

**University College London Hospital & University College London**
M Marks (PI), J S Brown (PI), N Ahwireng, B Bang, D Basire, R C Chambers, A Checkley, R Evans, M Heightman, T Hillman, J Hurst, J Jacob, S Janes, R Jastrub, M Lipman, S Logan, D Lomas, M Merida Morillas, A Pakzad, H Plant, J C Porter, K Roy, E Wall, B Williams, M Xu

**University Hospital Birmingham NHS Foundation Trust & University of Birmingham**
D Parekh (PI), N Ahmad Haider, C Atkin, R Baggott, M Bates, A Botkai, A Casey, B Cooper, J Dasgin, K Draxlbauer, N Gautam, J Hazeldine, T Hiwot, S Holden, K Isaacs, T Jackson, S Johnson, V Kamwa, D Lewis, J M Lord, S Madathil, C McGhee, K Mcgee, A Neal, A Newton Cox, J Nyaboko, D Parekh, Z Peterkin, H Qureshi, B Rangelov, L Ratcliffe, E Sapey, J Short, T Soulsby, R Steeds, J Stockley, Z Suleiman, T Thompson, M Ventura, S Walder, C Welch, D Wilson, S Yasmin, K P Yip

**University Hospitals of Derby and Burton**

P Beckett (PI) C Dickens, U Nanda

**University Hospitals of Leicester NHS Trust & University of Leicester**

C E Brightling (CI), R A Evans (PI), M Aljaroof, N Armstrong, H Arnold, H Aung, M Bakali, M Bakau, M Baldwin, M Bingham, M Bourne, C Bourne, N Brunskill, P Cairns, L Carr, A Charalambou, C Christie, M J Davies, S Diver, S Edwards, C Edwardson, O Elneima, H Evans, J Finch, S Glover, N Goodman, B Gootpu, N J Greening, B Guillen-Guio, K Hadley, P Haldar, B Hargadon, V C Harris, L Houchen-Wolloff, W Ibrahim, L Ingram, K Khunti, A Lea, D Lee, D Lozano-Rojas, G P McCann, H J C McAuley, P McCourt, T Mcnally, G Mills, A Moss, W Monteiro, K Ntotsis, M Pareek, S Parker, A Rowland, A Prickett, I N Qureshi, R Russell, N Samani, M Sereno, M Sharma, A Shikotra, S Siddiqui, A Singapuri, S J Singh, J Skeemer, M Soares, E Stringer, T Thornton, M Tobin, E Turner, L V Wain, T J C Ward, F Woodhead, J Wormleighton, T Yates, A Yousuf,

**University Hospital Southampton NHS Foundation Trust & University of Southampton**
M G Jones (PI), C Childs, R Djukanovic, S Fletcher, M Harvey, E Marouzet, B Marshall, R Samuel, T Sass, T Wallis, H Wheeler

**Whittington Health NHS**
R Dharmagunawardena (PI), E Bright, P Crisp, M Stern

**Wirral University Teaching Hospital**

A Wight (PI), L Bailey, A Reddington

**Wrightington Wigan and Leigh NHS trust**

A Ashish (PI), J Cooper, E Robinson

**Yeovil District Hospital NHS Foundation Trust**

A Broadley (PI)

**York & Scarborough NHS Foundation Trust**

K Howard (PI), L Barman, C Brookes, K Elliott. L Griffiths, Z Guy, D Ionita, H Redfearn, C Sarginson
A Turnbull

**Health and Care Research Wales**

Y Ellis

**London School of Hygiene & Tropical Medicine (LSHTM)**
M Marks, A Briggs

**NIHR Office for Clinical Research Infrastructure**

K Holmes

**Patient Public Involvement Leads**

Asthma UK and British Lung Foundation Partnership - K Poinasamy, S Walker

**Royal Surrey NHS Foundation Trust**
M Halling-Brown

**South London and Maudsley NHS Foundation Trust & Kings College London**
G Breen, M Hotopf

**Swansea University & Swansea Welsh Network**

K Lewis, N Williams

#### Appendix B: Detail on Statistical Methods

##### EQ5D-5L Utility Index Scores

EQ5D utility scores ($U$) (higher score indicates a better quality of life) were transformed to disutility scores ($D$) (higher score indicates worse quality of life) using a simple linear transformation $D = 1 - U$ to give a distribution of values with a right skew which is more easily amenable for statistical analysis. Then, in order to examine the potential effects of the health care pathway covariates on the EQ5D disutility at 12-months while controlling for potential differences in baseline demographic and clinical risk factors, a generalised linear mixed model (GLMM) model of the form:

$$ln\ (E[\mathrm{D}])\ = \mathrm{X\beta} + \mathrm{Y\gamma} + \upsilon$$

was constructed, assuming a gamma distribution of disutility and a long link. $D$ is the EQ5D disutility index score, X are the covariates to be adjusted for, with coefficients $\beta$, $Y$ are the indicators of health care pathway, with coefficients $\gamma$, and $\upsilon$ is a random intercept at the level of the hospital. We run our statistical analysis in R and use the *glmmTMB* package (Brooks et al, 2017).

##### Healthcare Resource Use and Associated Costs

In order to examine the potential effect of the health care pathway covariates on the accumulated healthcare costs at 12 months post-discharge while controlling for potential differences in baseline demographic and clinical risk factors we construct a GLM model for cost with a Gamma distribution and log link:

$$ln\ (E[\mathrm{C}])\ = \mathrm{X\beta} + \mathrm{Y\gamma} + \upsilon$$

where C is the health care costs and the right hand side is as previously specified for disutility.

#### Appendix C – Supplementary Tables

**Table S1**
**Healthcare Resource items and associated unit costs**

| Healthcare Resource | Unit Cost (£) | Source |
|---|---|---|
| *Rehabilitation* | | |
| - Initial assessment | 123 | NSHSC |
| - 10 sessions | 930 | NSHSC |
| *Mental Health Services* | | |
| - Post ICU psychology service | 507 | NSHSC |
| - Psychiatric liaison service (1.3 sessions) | 352 | NSHSC |
| - Improving Access to Psychological Therapies (7.5 sessions) | 990 | PSSRU |
| - Acute Hospital Clinical Health Psychology service | 155 | NSHSC |
| - Community Mental Health Team (adult) (8.7 sessions) | 2,047 | NSHSC |
| - Community Mental Health Team (older adult) (8.7 sessions) | 2,056 | NSHSC |
| - Mental health Crisis Resolution and Home Treatment service | 354 | NSHSC |
| - In patient mental health service | 632 | NSHSC |
| - Counselling services (Third sector or primary care based) | 279 | NSHSC |
| - Private providers of psychological therapy | 38 | PSSRU |
| *Basic Investigations* | | |
| - Chest X-Ray | 32 | SoECAT |
| - Electrocardiogram | 27 | SoECAT |
| *Blood tests* | | |
| - FBC | 12 | SoECAT |
| - U&Es | 8 | SoECAT |
| - eGFR | 12 | median |
| - LFTs | 12 | SoECAT |
| - CRP | 11 | SoECAT |
| - Bone profile | 8 | SoECAT |
| - 25-Hydroxyvitamin D (25-OH) | 12 | median |
| - BNP/Pro-NT BNP | 35 | SoECAT |
| - Troponin | 23 | SoECAT |
| - D Dimer | 15 | SoECAT |
| - Fibrinogen | 10 | SoECAT |
| - INR | 11 | SoECAT |
| - Ferritin | 11 | SoECAT |
| - HbA1C | 8 | SoECAT |
| - Lipid profile (non-fasting) | 12 | SoECAT |
| - SARS-Cov-2 Serology | 46 | SoECAT |
| *Respiratory Investigations and Procedures* | | |
| - Spirometry | 22 | SoECAT |
| - Full pulmonary function tests | 72 | NSHSC |
| - Fractional exhaled nitric oxide | 72 | NSHSC |
| - Nijmegen questionnaire | 22 | SoECAT |

| | | |
|---|---|---|
| - Incremental shuttle walk test (ISWT) | 207 | NSHSC |
| - Six-minute walk test | 207 | NSHSC |
| - Cardio-pulmonary exercise test | 234 | NSHSC |
| - CT chest with contrast | 238 | SoECAT |
| - HRCT | 157 | SoECAT |
| - CTPA | 157 | SoECAT |
| - Pulmonary perfusion scan | 269 | NSHSC |
| *Cardiac Investigations and Procedures* | | |
| - Cardiac MRI | 374 | SoECAT |
| - Echocardiogram | 27 | SoECAT |
| - 24/48 hrs tape | 102 | SoECAT |
| - Right heart catheterisation | 258 | NSHSC |
| - Coronary angiogram/CT angiogram | 157 | SoECAT |
| - Cardiac perfusion scan | 344 | NSHSC |
| *Haematological Investigations and Procedures* | | |
| - Thyroid function tests | 30 | SoECAT |
| - Blood borne viruses screen | 30 | median |
| - Haematinics | 30 | median |
| *Renal Investigations and Procedures* | | |
| - USS abdomen | 59 | SoECAT |
| - USS Kidneys | 59 | SoECAT |
| - CT abdomen/pelvis | 157 | SoECAT |
| - Urinalysis | 22 | SoECAT |
| - Urine albumin: creatinine ratio | 22 | SoECAT |
| - Urine protein: creatinine ratio | 22 | SoECAT |
| *Immunological Investigations and Procedures* | | |
| - Complements factors | 23 | SoECAT |
| - Immunoglobulins | 23 | SoECAT |
| - Rheumatoid factor/Anti-CCP | 23 | SoECAT |
| *Neurology/Mental health Investigations and Procedures* | | |
| - CT brain | 157 | SoECAT |
| - MRI brain | 374 | SoECAT |
| - Nerve conduction studies | 100 | NSHSC |
| - Sleep Condition Indicator | 22 | SoECAT |
| *Emergency visit ITU/HDU admission* | | |
| - Short stay | 827 | PSSRU |
| - Long stay (>= 2 days) | 3,627 | PSSRU |
| Outpatient visit | 137 | PSSRU |
| Contacts with GP for a clinical review | 39 | PSSRU |

NSHSC = National Schedule of NHS Costs; PSSRU = Personal Social Services Research Unit; SoECAT = Schedule of Events Cost Attribution Tool; FBC - Full Blood Count; URES - Urea and Electrolytes; eGFR - Estimated Glomerular Filtration Rate; LFTs - Liver Function Tests; CRP - C-Reactive Protein; Pro-NT BNP - N-terminal pro b-type Natriuretic Peptide; INR - International Normalised Ratio; HbA1c - Haemoglobin A1c; SARS-CoV-2 - Severe Acute Respiratory Syndrome Coronavirus 2; ISWT - Incremental Shuttle Walk Test; CT – Computed Tomography; HRCT - High-Resolution Computed Tomography; CTPA - Computed Tomography Pulmonary Angiography; MRI - Magnetic Resonance Imaging; USS - Ultrasound Scan; Anti-CCP - Anti-Cyclic Citrullinated Peptide; ITU - Intensive Therapy Unit; HDU - High Dependency Unit; ; GP - General Practitioner.

**Table S2**
**Gamma distributed log-link GLM for EQ5D disutility**

| | | | | 95% CI | |
|---|---|---|---|---|---|
| | Estimate | SE | p-value | Lower | Upper |
| Intercept | 0.208 | 0.208 | 0.317 | -0.200 | 0.617 |
| Female | 0.237 | 0.062 | 0.000 | 0.115 | 0.359 |
| Pre-COVID Utility Index Summary Score | -2.136 | 0.156 | 0.000 | -2.441 | -1.830 |
| Index of Multiple Deprivation quintile | | | | | |
| - 2 | 0.113 | 0.088 | 0.195 | -0.058 | 0.285 |
| - 3 | 0.060 | 0.092 | 0.510 | -0.119 | 0.240 |
| - 4 | -0.007 | 0.093 | 0.937 | -0.190 | 0.175 |
| - 5 - least deprived | -0.318 | 0.091 | 0.000 | -0.496 | -0.140 |
| Age at admission (years) | | | | | |
| - <30 | -0.173 | 0.254 | 0.496 | -0.672 | 0.325 |
| - 30–39 | -0.212 | 0.135 | 0.117 | -0.478 | 0.053 |
| - 40–49 | -0.105 | 0.100 | 0.294 | -0.301 | 0.091 |
| - 60–69 | -0.109 | 0.073 | 0.138 | -0.252 | 0.035 |
| - 70–79 | -0.252 | 0.093 | 0.007 | -0.435 | -0.070 |
| - 80+ | 0.093 | 0.172 | 0.587 | -0.244 | 0.431 |
| WHO respiratory support class | | | | | |
| - 5 | -0.138 | 0.087 | 0.111 | -0.308 | 0.032 |
| - 6 | -0.069 | 0.099 | 0.487 | -0.263 | 0.125 |
| - 7–9 | 0.370 | 0.100 | 0.000 | 0.175 | 0.566 |
| Baseline comorbidities | | | | | |
| - Cardiac | 0.065 | 0.064 | 0.310 | -0.060 | 0.189 |
| - Respiratory | 0.180 | 0.065 | 0.005 | 0.053 | 0.307 |
| - Gastrointestinal | 0.055 | 0.085 | 0.519 | -0.112 | 0.221 |
| - Neurological and psychiatric | 0.281 | 0.080 | 0.000 | 0.123 | 0.438 |
| - Rheumatological | 0.249 | 0.092 | 0.007 | 0.069 | 0.430 |
| - Metabolic/Endocrine/Renal | -0.019 | 0.091 | 0.832 | -0.198 | 0.159 |
| - Malignancy/Haematological | 0.074 | 0.122 | 0.544 | -0.165 | 0.313 |
| Ethnicity | | | | | |
| - South Asian | -0.115 | 0.109 | 0.295 | -0.329 | 0.100 |
| - Black | -0.101 | 0.113 | 0.371 | -0.323 | 0.121 |
| - Mixed | -0.190 | 0.201 | 0.344 | -0.583 | 0.203 |
| - Other | -0.151 | 0.156 | 0.332 | -0.456 | 0.154 |
| BMI >= 30kg/$m^2$ | 0.195 | 0.060 | 0.001 | 0.078 | 0.312 |
| Days since discharge | 0.000 | 0.000 | 0.583 | 0.000 | 0.000 |
| Healthcare pathway | | | | | |
| - Access and assessment | -0.223 | 0.097 | 0.022 | -0.414 | -0.032 |
| - Rehabilitation services | -0.178 | 0.065 | 0.006 | -0.306 | -0.051 |
| - Mental health services | 0.066 | 0.063 | 0.293 | -0.057 | 0.189 |
| - All patients offered services | 0.069 | 0.067 | 0.305 | -0.063 | 0.200 |

**Table S3**
**Gamma distributed log-link GLM for healthcare costs in 12-months post hospitalisation**

| | Estimate | SE | p-value | 95% CI Lower | 95% CI Upper |
|---|---|---|---|---|---|
| Intercept | 7.156 | 0.389 | 0.000 | 6.393 | 7.918 |
| Female | 0.236 | 0.079 | 0.003 | 0.082 | 0.390 |
| Pre-COVID Utility Index Summary Score | -0.417 | 0.199 | 0.036 | -0.807 | -0.028 |
| Index of Multiple Deprivation quintile | | | | | |
| - 2 | -0.102 | 0.112 | 0.362 | -0.321 | 0.117 |
| - 3 | -0.274 | 0.116 | 0.018 | -0.501 | -0.047 |
| - 4 | -0.339 | 0.116 | 0.003 | -0.566 | -0.112 |
| - 5 - least deprived | -0.350 | 0.117 | 0.003 | -0.580 | -0.120 |
| Age at admission (years) | | | | | |
| - <30 | -0.165 | 0.290 | 0.571 | -0.734 | 0.405 |
| - 30–39 | -0.361 | 0.167 | 0.030 | -0.688 | -0.035 |
| - 40–49 | -0.057 | 0.123 | 0.646 | -0.298 | 0.185 |
| - 60–69 | -0.232 | 0.089 | 0.010 | -0.407 | -0.057 |
| - 70–79 | -0.016 | 0.116 | 0.892 | -0.244 | 0.212 |
| - 80+ | 0.037 | 0.208 | 0.857 | -0.370 | 0.445 |
| WHO respiratory support class | | | | | |
| - 5 | -0.237 | 0.110 | 0.032 | -0.453 | -0.021 |
| - 6 | -0.050 | 0.122 | 0.682 | -0.288 | 0.189 |
| - 7–9 | 0.510 | 0.128 | 0.000 | 0.259 | 0.761 |
| Baseline comorbidities | | | | | |
| - Cardiac | 0.008 | 0.077 | 0.918 | -0.142 | 0.158 |
| - Respiratory | 0.520 | 0.080 | 0.000 | 0.363 | 0.678 |
| - Gastrointestinal | 0.096 | 0.106 | 0.364 | -0.111 | 0.303 |
| - Neurological and psychiatric | 0.189 | 0.101 | 0.060 | -0.008 | 0.387 |
| - Rheumatological | -0.089 | 0.121 | 0.464 | -0.326 | 0.149 |
| - Metabolic/Endocrine/Renal | 0.102 | 0.115 | 0.374 | -0.124 | 0.328 |
| - Malignancy/Haematological | 0.307 | 0.148 | 0.038 | 0.017 | 0.597 |
| Ethnicity | | | | | |
| - South Asian | -0.247 | 0.145 | 0.089 | -0.532 | 0.037 |
| - Black | -0.037 | 0.242 | 0.878 | -0.511 | 0.436 |
| - Mixed | -0.214 | 0.191 | 0.264 | -0.589 | 0.161 |
| - Other | -0.142 | 0.077 | 0.063 | -0.293 | 0.008 |
| BMI >= 30kg/$m^2$ | -0.239 | 0.129 | 0.065 | -0.493 | 0.014 |
| Healthcare pathway | | | | | |
| - Access and assessment | -0.049 | 0.358 | 0.892 | -0.750 | 0.653 |
| - Rehabilitation services | 0.034 | 0.299 | 0.909 | -0.551 | 0.619 |
| - Mental health services | -0.122 | 0.301 | 0.684 | -0.712 | 0.467 |
| - All patients offered service | 0.312 | 0.310 | 0.313 | -0.295 | 0.919 |

**Table S4**
**Gamma distributed log-link GLM for EQ5D (weighted)**

| | | | | 95% CI | |
|---|---|---|---|---|---|
| | Estimate | SE | p-value | Lower | Upper |
| Intercept | 0.208 | 0.208 | 0.317 | -0.200 | 0.617 |
| Female | 0.237 | 0.062 | 0.000 | 0.115 | 0.359 |
| Pre-COVID Utility Index Summary Score | -2.136 | 0.156 | 0.000 | -2.441 | -1.830 |
| Index of Multiple Deprivation quintile | | | | | |
| - 2 | 0.113 | 0.088 | 0.195 | -0.058 | 0.285 |
| - 3 | 0.060 | 0.092 | 0.510 | -0.119 | 0.240 |
| - 4 | -0.007 | 0.093 | 0.937 | -0.190 | 0.175 |
| - 5 - least deprived | -0.318 | 0.091 | 0.000 | -0.496 | -0.140 |
| Age at admission (years) | | | | | |
| - <30 | -0.173 | 0.254 | 0.496 | -0.672 | 0.325 |
| - 30–39 | -0.212 | 0.135 | 0.117 | -0.478 | 0.053 |
| - 40–49 | -0.105 | 0.100 | 0.294 | -0.301 | 0.091 |
| - 60–69 | -0.109 | 0.073 | 0.138 | -0.252 | 0.035 |
| - 70–79 | -0.252 | 0.093 | 0.007 | -0.435 | -0.070 |
| - 80+ | 0.093 | 0.172 | 0.587 | -0.244 | 0.431 |
| WHO respiratory support class | | | | | |
| - 5 | -0.138 | 0.087 | 0.111 | -0.308 | 0.032 |
| - 6 | -0.069 | 0.099 | 0.487 | -0.263 | 0.125 |
| - 7–9 | 0.370 | 0.100 | 0.000 | 0.175 | 0.566 |
| Baseline comorbidities | | | | | |
| - Cardiac | 0.065 | 0.064 | 0.310 | -0.060 | 0.189 |
| - Respiratory | 0.180 | 0.065 | 0.005 | 0.053 | 0.307 |
| - Gastrointestinal | 0.055 | 0.085 | 0.519 | -0.112 | 0.221 |
| - Neurological and psychiatric | 0.281 | 0.080 | 0.000 | 0.123 | 0.438 |
| - Rheumatological | 0.249 | 0.092 | 0.007 | 0.069 | 0.430 |
| - Metabolic/Endocrine/Renal | -0.019 | 0.091 | 0.832 | -0.198 | 0.159 |
| - Malignancy/Haematological | 0.074 | 0.122 | 0.544 | -0.165 | 0.313 |
| Ethnicity | | | | | |
| - South Asian | -0.115 | 0.109 | 0.295 | -0.329 | 0.100 |
| - Black | -0.101 | 0.113 | 0.371 | -0.323 | 0.121 |
| - Mixed | -0.190 | 0.201 | 0.344 | -0.583 | 0.203 |
| - Other | -0.151 | 0.156 | 0.332 | -0.456 | 0.154 |
| BMI >= 30kg/$m^2$ | 0.195 | 0.060 | 0.001 | 0.078 | 0.312 |
| Days since discharge | 0.000 | 0.000 | 0.583 | 0.000 | 0.000 |
| Healthcare pathway | | | | | |
| - Access and assessment | -0.223 | 0.097 | 0.022 | -0.414 | -0.032 |
| - Rehabilitation services | -0.178 | 0.065 | 0.006 | -0.306 | -0.051 |
| - Mental health services | 0.066 | 0.063 | 0.293 | -0.057 | 0.189 |
| - All patients offered services | 0.069 | 0.067 | 0.305 | -0.063 | 0.200 |

**Table S5**
**Gamma distributed log-link GLM for healthcare costs in 12-months post hospitalisation (weighted)**

| | | | | 95% CI | |
|---|---|---|---|---|---|
| | Estimate | SE | p-value | Lower | Upper |
| Intercept | 6.822 | 0.400 | 0.000 | 6.039 | 7.605 |
| Female | 0.083 | 0.076 | 0.274 | -0.066 | 0.232 |
| Pre-COVID Utility Index Summary Score | -0.348 | 0.177 | 0.049 | -0.695 | -0.002 |
| Index of Multiple Deprivation quintile | | | | | |
| - 2 | -0.065 | 0.096 | 0.497 | -0.254 | 0.123 |
| - 3 | 0.039 | 0.109 | 0.718 | -0.174 | 0.252 |
| - 4 | -0.001 | 0.111 | 0.990 | -0.219 | 0.216 |
| - 5 - least deprived | -0.033 | 0.120 | 0.786 | -0.267 | 0.202 |
| Age at admission (years) | | | | | |
| - <30 | -0.734 | 0.142 | 0.000 | -1.013 | -0.456 |
| - 30–39 | -0.398 | 0.142 | 0.005 | -0.676 | -0.119 |
| - 40–49 | 0.124 | 0.132 | 0.346 | -0.134 | 0.383 |
| - 60–69 | -0.234 | 0.119 | 0.050 | -0.467 | 0.000 |
| - 70–79 | -0.108 | 0.120 | 0.371 | -0.343 | 0.128 |
| - 80+ | 0.076 | 0.133 | 0.566 | -0.184 | 0.337 |
| WHO respiratory support class | | | | | |
| - 5 | -0.002 | 0.085 | 0.979 | -0.170 | 0.165 |
| - 6 | 0.166 | 0.104 | 0.111 | -0.038 | 0.371 |
| - 7–9 | 0.749 | 0.200 | 0.000 | 0.357 | 1.141 |
| Baseline comorbidities | | | | | |
| - Cardiac | -0.116 | 0.083 | 0.165 | -0.279 | 0.048 |
| - Respiratory | 0.691 | 0.081 | 0.000 | 0.533 | 0.850 |
| - Gastrointestinal | 0.108 | 0.100 | 0.279 | -0.088 | 0.304 |
| - Neurological and psychiatric | -0.013 | 0.104 | 0.899 | -0.218 | 0.191 |
| - Rheumatological | 0.001 | 0.119 | 0.991 | -0.231 | 0.234 |
| - Metabolic/Endocrine/Renal | 0.039 | 0.111 | 0.728 | -0.180 | 0.257 |
| - Malignancy/Haematological | 0.342 | 0.153 | 0.025 | 0.043 | 0.641 |
| Ethnicity | | | | | |
| - South Asian | -0.385 | 0.112 | 0.001 | -0.604 | -0.166 |
| - Black | 0.231 | 0.136 | 0.089 | -0.035 | 0.497 |
| - Mixed | -0.082 | 0.234 | 0.727 | -0.539 | 0.376 |
| - Other | -0.033 | 0.226 | 0.884 | -0.477 | 0.410 |
| BMI >= 30kg/m$^2$ | 0.009 | 0.073 | 0.896 | -0.133 | 0.152 |
| Healthcare pathway | | | | | |
| - Access and assessment | 0.036 | 0.394 | 0.928 | -0.736 | 0.807 |
| - Rehabilitation services | -0.181 | 0.315 | 0.565 | -0.798 | 0.436 |
| - Mental health services | -0.228 | 0.317 | 0.473 | -0.850 | 0.394 |
| - All patients offered service | 0.415 | 0.333 | 0.212 | -0.237 | 1.066 |
